# Brain network mechanisms of visual perceptual organization in schizophrenia and bipolar disorder

**DOI:** 10.1101/2022.01.26.22269913

**Authors:** Brian P. Keane, Bart Krekelberg, Ravi D. Mill, Steven M. Silverstein, Judy L. Thompson, Megan Serody, Deanna M. Barch, Michael W. Cole

## Abstract

Visual shape completion is a canonical perceptual organization process that integrates spatially distributed edge information into unified representations of objects. People with schizophrenia show difficulty in discriminating completed shapes but the brain networks and functional connections underlying this perceptual difference remain poorly understood. Also unclear is whether similar neural differences arise in bipolar disorder or vary across the schizo-bipolar spectrum. To address these topics, we scanned (fMRI) people with schizophrenia, bipolar disorder, or no psychiatric illness during rest and during a task in which they discriminated configurations that formed or failed to form completed shapes (illusory and fragmented condition, respectively). Multivariate pattern differences were identified on the cortical surface using 360 predefined parcels and 12 functional networks composed of such parcels. Brain activity flow mapping was used to evaluate the likely involvement of resting-state connections for shape completion. Illusory/fragmented task activation differences (“modulations”) in the dorsal attention network (DAN) could distinguish people with schizophrenia (AUCs>.85) and could trans-diagnostically predict cognitive disorganization severity. Activity flow over functional connections from the DAN could predict secondary visual network modulations in each group, except among those with schizophrenia. The secondary visual network was strongly and similarly modulated in each subject group. Task modulations were dispersed over a larger number of networks in patients compared to controls. In summary, abnormal DAN activity emerges during perceptual organization in schizophrenia and may be related to improper attention-related feedback into secondary visual areas. Patients with either disorder may compensate for abnormal perception by relying upon non-visual networks.

## 1. Introduction

Schizophrenia (SZ) is a debilitating disorder charactered by delusions, hallucinations, disorganized thought and a decline in social/occupational functioning. The disorder also adversely impacts aspects of visual perceptual organization (Silverstein and Keane, 2011) and, in particular, visual shape completion, which builds shape representations from co-aligned step-edge elements (Keane et al., 2019). Why might this be? The question is important, first, because shape completion plays an absolutely fundamental role for normal seeing (Keane, 2018), recovering object shape, size, and number within camouflaged or cluttered distal environments. Understanding subtle impairments in shape completion and related perceptual organization processes (Silverstein and Keane, 2011) could help us understand, for example, why individuals with the disorder have uncomfortable sensations of sensory flooding (Bunney et al., 1999) or why they have poorer overall day-to-day visual functioning (Shoham et al., 2020). Another reason to investigate the brain-basis of shape completion in schizophrenia is that the underlying mechanisms are already partly understood. Extensive investigations in human and non-human primates using single-unit recording, TMS, EEG, and fMRI, have all shown that shape completion relies upon a mid-level visual regions such as lateral occipital cortex and V4, as well recursive interactions with V1 and V2 (Cox et al., 2013; Murray et al., 2006). A recent brain network analysis, described further below, has additionally revealed that shape completion activates a sparse but densely interconnected coalition of regions that is seated in the secondary visual network and that incorporates pieces of at least four other networks (Keane et al., 2021a). Patient neuroimaging findings can thus be situated within this existing literature.

We postulate on the basis of past work that SZ patients form illusory contours at initial stages of processing but do not properly use such contours at later, conceptual stages. As evidence, in a visual evoked potential study, when participants discriminated configurations that formed or failed to form illusory shapes, there was an intact illusory contour formation waveform over lateral occipital regions at 106-194 ms post-stimulus onset (see also, Wynn et al., 2015) and an increased, possibly compensatory, “closure negativity” waveform across frontal regions at 240-400 ms. In a methodologically similar EEG study, when subjects discriminated illusory square from non-illusory (fragmented) stimuli, SZ patients exhibited a unique response-locked high-gamma oscillation—with a fronto-temporal topography—at a relatively late processing stage (100 ms before button-press) (Spencer and Ghorashi, 2014). In a psychophysical study, schizophrenia patients reacted normally to distractor lines placed near illusory contours, suggesting intact illusory contour formation, but were overall poor at discriminating illusory shapes, suggesting a lessened ability to notice and use contours (Keane et al., 2021b; 2014). In a non-clinical psychophysical study, when certain participants were cognitively biased through instructional templates and verbal instructions to see the inducing pac-man edges as disconnected, these individuals performed as if they had schizophrenia; that is, they normally reacted to distractor lines, normally discriminated non-illusory stimuli (which fail to form illusory contours), but poorly discriminated illusory shapes (Keane et al., 2012). The foregoing results, taken together, suggest that patients’ visual networks may operate relatively normally during shape completion but that higher-order cognitive networks may not. A purpose of the present investigation is to verify this assertion with functional MRI. A second goal was to consider the neural basis of shape completion in bipolar disorder. Bipolar disorder was considered, first, because it offers an important foil for schizophrenia. Over 40% of bipolar disorder patients take anti-psychotic medications (Rhee et al., 2020), over half report at least one lifetime psychotic symptom (Dunayevich and Keck, 2000), both are associated with chronic medical problems and past substance abuse history (Cassidy et al., 2001; Dixon, 1999), and there is genetic overlap between the two (Lichtenstein et al., 2009). Therefore, establishing group differences would more convincingly demonstrate specificity to schizophrenia. Moreover, understanding visual disturbances in bipolar disorder is important in its own right. Despite being twice as prevalent as schizophrenia (American Psychiatric Association, 2013), a PubMed title/abstract search yielded 5% as much literature on “visual perception” (Search date: October 27^th^, 2021). We had no hypotheses regarding which networks would be affected, given the dearth of data on the subject.

A third goal was to move beyond the traditional DSM-5 nosology and to probe for neural signatures that might be shared between disorders or that might depend on factors that cut across the schizo-bipolar spectrum (Kozak and Cuthbert, 2016). Based on past work, we expected to find neural signatures linked to cognitive disorganization, a cardinal symptom of schizophrenia (Keane et al., 2019; Spencer and Ghorashi, 2014; Spencer et al., 2004). This link was expected to emerge within a brain network that was differentially modulated in schizophrenia.

A final and more exploratory question pertained to the extent to which top-down feedback might influence activity in the secondary visual network during shape completion, given the critical role of this network for shape completion (Keane et al., 2021a). We have provided evidence that—in healthy controls—the dorsal attention network (DAN) plausibly acts as a bridge to the secondary visual network and we have supported this claim with a “brain activity flow” modeling procedure (“ActFlow”) in which task activations and resting-state functional connections from the DAN could be used to model task activations in the secondary visual network (Cole et al., 2016; Keane et al., 2021a). There are reasons to think that such feedback should be weak in schizophrenia. For example, dynamic causal modelling has shown that—during a depth inversion illusion task—SZ patients exhibited poor top-down feedback from the intraparietal sulcus (in the dorsal attention network) to the lateral occipital cortex (Dima et al., 2010) but normal feedforward activity between the same regions. Other studies have shown intact subliminal processing of masked words or digits, indicating that much of bottom-up processing may be preserved (Berkovitch et al., 2017). If top-down feedback is indeed impaired in schizophrenia, then the above-described modeling effort should not be successful in schizophrenia patients.

To consider the above questions, we scanned 16 schizophrenia (SZ) participants and 15 people with bipolar disorder (BP) during rest and during a task in which they discriminated pac-man configurations that either formed or failed to form visually completed shapes (illusory and fragmented condition, respectively) (Ringach and Shapley, 1996). These results were compared to healthy control data (n=20) that were already reported in an earlier study (Keane et al., 2021a). Our sample sizes were obviously not large. However, because our analyses were conducted on networks rather than individual regions, we were able to bypass massive multiple comparison correction and pool over functionally related cortical areas to magnify any possible group effects (Cremers et al., 2017; Ji et al., 2019; Noble et al., 2021). Similar to past studies, shape completion was operationalized as the difference in performance or activation between the illusory/fragmented conditions (Keane et al., 2021a; 2019). This so-called “fat/thin” task was chosen because it has been used extensively to investigate shape completion via psychophysics, fMRI, EEG, and TMS (Maertens and Pollmann, 2005; Murray et al., 2006; Pillow and Rubin, 2002; Wokke et al., 2013) and because it has also been used to demonstrate shape completion deficits in past behavioral work in schizophrenia (Keane et al., 2019). The resting-state scan data allowed us to compute the resting-state functional connectivity (RSFC) matrix between all pairs of regions, which in turn allowed us to model top-down feedback into the secondary visual network.

The above-stated hypotheses were tested in five steps. First, in each group, we divided the parcels into 12 functional networks (Ji et al., 2019) and quantified each network’s contribution to shape completion by applying MVPA to parcel-wise illusory and fragmented task activations. The networks that were of special interest were those traditionally associated with cognitive control (frontoparietal, dorsal attention, default mode, cingulo-opercular) and visual perceptual organization (secondary visual network). In the next step, for each pair of subject groups, we identified the networks that were differentially modulated by applying MVPA to parcel-wise task activation *differences* (illusory-fragmented). Third, we employed cross-validation and permutation testing to consider whether task activation differences within the DAN (whose relevance was established in steps 1 and 2) could predict cognitive disorganization severity. In a fourth preparatory step described in the Supplementary Material, we computed the resting-state functional connectomes (RSFC matrices) and demonstrated the likely utility of these functional connections for shape completion via ActFlow. Finally, again using ActFlow, we determined which network contained the most informative resting-state connections for inferring differential task activity in the secondary visual network (whose relevance was established in Step 1). Brain activity flow mapping allowed us to determine whether the DAN (and other networks) could model activity in the secondary visual network in each group, which in turn provided clues on how the groups might differ on top-down modulation during shape completion.

## 2. Materials and Methods

### 2.1. Participants

The sample consisted of 20 healthy controls (HCs), 15 people with bipolar disorder (BPs; type I, type II, and 1 unspecified), and 16 people with schizophrenia including one with schizoaffective disorder (SZs; See Table 1). The control data were separately published to establish the normal brain network mechanisms of shape completion and to set the stage for patient comparisons (Keane et al., 2021a). One control and one bipolar participant lacked resting-state data but were still included in the task analyses. Patients were recruited from the Newark and Piscataway outpatient and partial hospital clinics at Rutgers University Behavioral Health Care (with one exception being a schizophrenia patient from the Nathan Kline Institute in Orangeburg NY). Controls were recruited from the same metropolitan areas. To prevent exaggerated group differences in IQ and education, controls without four-year college degrees were preferentially recruited. As can be seen from Table 1, groups did not differ on age, education (self/parental), smoking habits, handedness or gender; the patient groups did not differ on illness duration, olanzapine/imipramine equivalents or current/premorbid functioning.

**Table 1.**
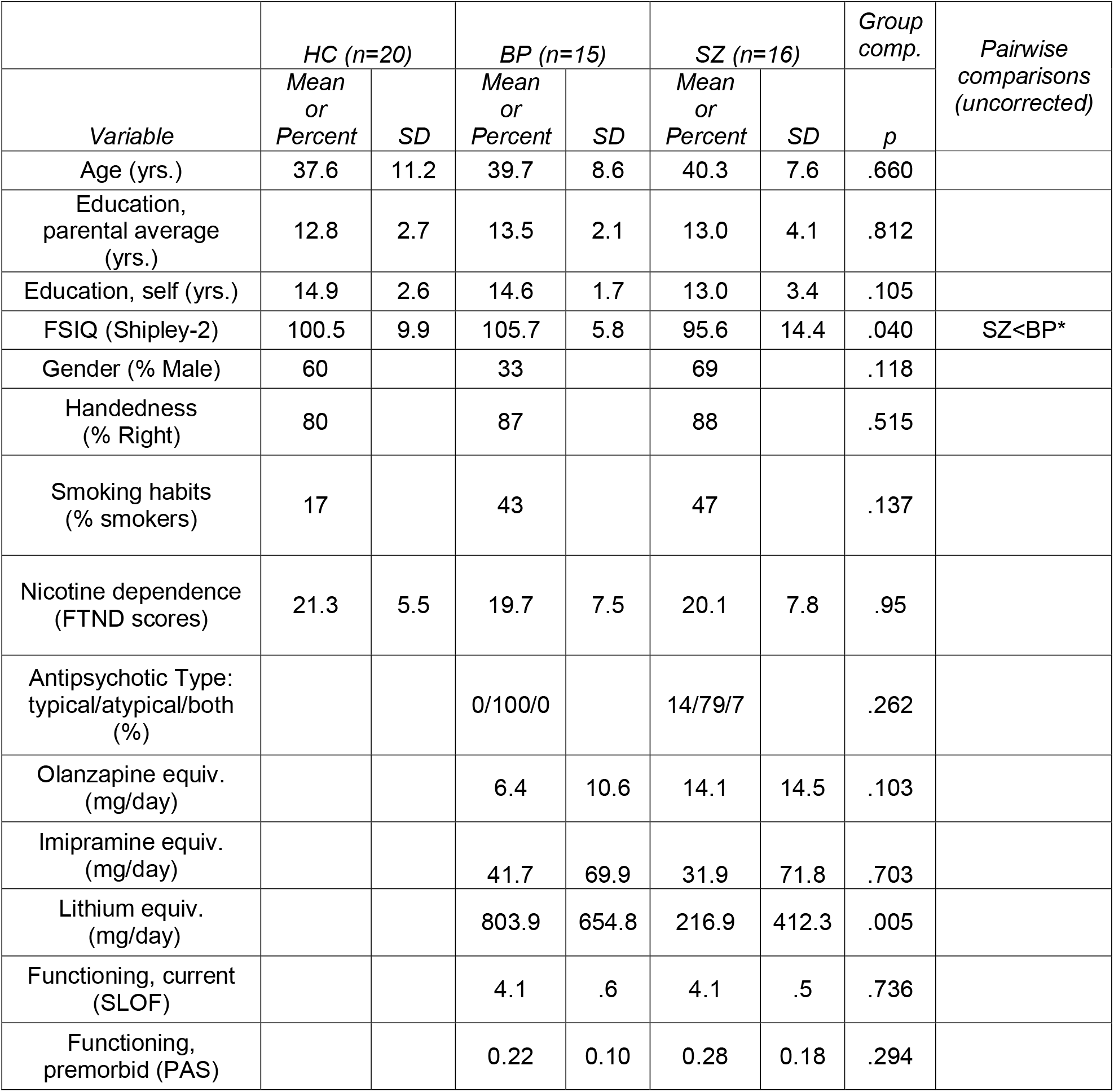
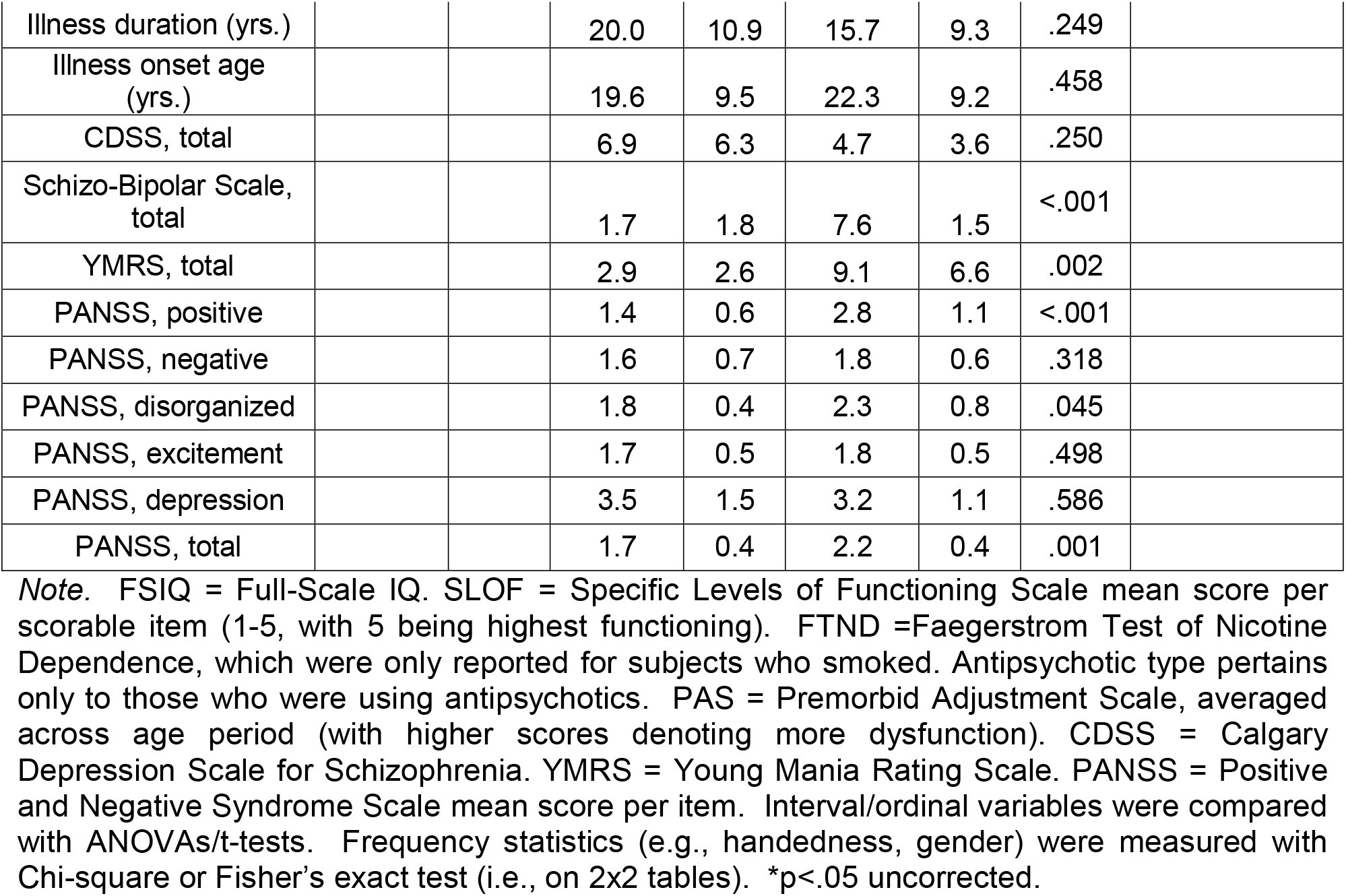
Demographic and clinical characteristics of participants.

The inclusion/exclusion criteria for all subjects were: (1) age 21-55; (2) no electroconvulsive therapy in the past 8 weeks; (3) no neurological or pervasive developmental disorders; (4) no recent substance use disorder (i.e., participants must not have satisfied more than one of the 11 Criterion A symptoms of DSM-5 substance use disorder in the last three months); (5) no positive urine toxicology screen or breathalyzer test on any day of testing, including THC; (6) no brain injury due to accident or illness (e.g., stroke or brain tumor) and no accompanying loss of consciousness for more than 10 minutes; (7) no amblyopia (as assessed by informal observation and self-report); (8) visual acuity of 20/32 or better (with corrective lenses if necessary); (9) the ability to understand English and provide written informed consent; (10) no scanner related contraindications (no claustrophobia, an ability to fit within the scanner bed, and no non-removable ferromagnetic material on or within the body); 11) no intellectual impairment (IQ<70) as assessed with brief vocabulary test (Shipley-2; see below). Additional criteria for controls were: (1) no DSM-5 diagnosis of past or current psychotic or mood disorders (including past mood episode); (2) no current psychotropic- or cognition-enhancing medication; (3) no first-degree relative with schizophrenia, schizoaffective, or bipolar disorder (as indicated by self-report). Additional criteria for patients were: (1) a DSM-5 diagnosis of schizophrenia, schizoaffective (depressive subtype), or bipolar disorder. Patients could not be in a manic state at the time of testing.

The participants in our study—while less numerous—were well-vetted and highly functioning, with few comorbidities. Of the consented HCs, 6 met DSM-5 criteria for an undisclosed past or current mood disorder (typically major depressive disorder), 1 could not reach a visual acuity of 20/32, 1 had amblyopia, 1 had scanner related contraindications, 4 tested positive for recreational or illicit substances, and 1 was excluded due to a safety concern. Of the consented BPs, 3 were excluded due to age (verified with ID), 6 had an IQ < 70, 2 could not reach a visual acuity of 20/32, 1 was excluded due to a head injury, 1 had scanner related contraindications, 1 had an alcohol use disorder, 2 tested positive for recreational or illicit substances, 1 had a neurological disorder, and 4 were excluded for multiple reasons. Of the consented SZs, 11 had an IQ < 70, 1 had amblyopia, 1 had scanner related contraindications, 2 had alcohol use disorders, 4 tested positive for recreational or illicit substances, and 2 were excluded for multiple reasons. The foregoing exclusions were in addition to those who were screened out before the consent, who ultimately received an inappropriate diagnosis, or who were disqualified for exhibiting COVID-19 symptoms or who had comorbidities that exacerbated the risks of COVID-19 (in the last year of recruitment).

Written informed consent was obtained from all subjects after explanation of the nature and possible consequences of participation. The study followed the tenets of the Declaration of Helsinki and was approved by the Rutgers University Institutional Review Board. All participants received monetary compensation and were naive to the study’s objectives.

### 2.2. Assessments

Psychiatric diagnosis was assessed with the Structured Clinical Interview for DSM-5 (28) and was assigned only after consulting detailed medical history and the SCID. All diagnoses were further considered during a weekly diagnostic consensus meeting. All clinical instruments were administered by a rater who had established reliability with raters in other ongoing studies (ICC > 0.8).

Intellectual functioning of all subjects was assessed with a brief vocabulary test that correlates highly (*r*=0.80) with WAIS-III full-scale IQ scores (Canivez and Watkins, 2010; W. C. Shipley et al., 2009, p. 65). Visual acuity was measured with a logarithmic visual acuity chart under fluorescent overhead lighting (viewing distance = 2 meters, lower limit =20/10), and in-house visual acuity correction was used for individuals without appropriate glasses or contacts. The Alere iCup Dx Drug Screen Cup was utilized to probe for the presence of recreational and illicit substances (i.e., THC, cocaine, methamphetamines, amphetamines, and opiates). The AlcoHawk Pro breathalyzer was administered to test for recent alcohol consumption. All included subjects tested negative for each test at the time of scanning. Nicotine use was assayed with the Faegerstrom Test for Nicotine Dependence (FTND) (Heatherton et al., 1991). Standardized medication dose equivalents (olanzapine, lithium, and imipramine equivalents) were determined for each patient using published tables (Bollini et al., 1999; Gardner et al., 2010) (Table 1).

The Positive and Negative Syndrome Scale (PANSS; Kay et al., 1987) was administered within two weeks of the scan and provided information about symptoms over the last two weeks. PANSS symptom scores were reported via a “consensus” 5-factor model, which was designed on the basis of 29 previous five-factor models (Wallwork et al., 2012). The disorganization score was the clinical variable of greatest interest, given its previously documented relation to shape completion (Keane et al., 2019).

To fully characterize the patient samples, we also administered several other symptom/functioning assessments. Depressive and manic symptoms were assessed with the Calgary Depression Scale in Schizophrenia (D. Addington et al., 1993) and the Young Mania Rating Scale, respectively (Young et al., 1978). The Specific Levels of Functioning Scale (SLOF) estimated day-to-day functioning in areas such as physical functioning, personal care, interpersonal relationships, social acceptability, activities, and work skills. The Premorbid Adjustment Scale (PAS; Cannon-Spoor et al., 1982) measured sociability, peer relationship quality, scholastic performance, school adaptation, and (where appropriate) social-sexual functioning up to 1 year before illness onset; this was done for childhood (up through age 11), early adolescence (ages 12-15), late adolescence (ages 16-18), and adulthood (ages 19 and above). In line with what others have done, the PAS General score was not included since it is reflective of functioning before and after illness onset (van Mastrigt and J. Addington, 2002). For individuals with schizophrenia, illness onset on the PAS was defined as when one or more positive symptoms first became noticeable or concerning to the patient. For individuals with bipolar disorder, illness onset was defined as the onset of the first mood episode (either manic or major depressive). Each patient’s position along the schizo-bipolar spectrum was assessed with the Schizo-Bipolar Scale (Keshavan et al., 2011). Higher scores indicated that a subject was more toward the pure ‘schizophrenia’ end of the spectrum.

### 2.3. Experimental Design and Statistical Analysis

#### 2.3.1. Stimulus and procedure

Participants performed a “fat/thin” shape discrimination task in which they indicated whether four pac-men formed a fat or thin shape (“illusory” condition) or whether four downward-facing pac-men were uniformly rotated left or right (“fragmented” condition) (see Fig. 1). The fragmented task is a suitable control in that it involves judging the lateral properties of the stimulus—just like the illusory condition—and in that it uses groupable elements (via common orientation, Beck, 1966). As described elsewhere (Keane et al., 2021), the two tasks shared most stimulus and procedural details (stimulus timing, pac-man features, spatial distribution, etc.) and therefore relied on many of the same processes (temporal attention, divided attention, visual working memory, etc.) (Keane et al., 2019). Perhaps because of these similarities, the tasks generate similar performance thresholds, reaction times, and accuracies, and are highly correlated behaviorally (Keane et al., 2021a; 2019), which is interesting since extremely similar visual tasks are often uncorrelated even within large samples (Grzeczkowski et al., 2017). In sum, by having employed a closely matched and already tested control condition, we were in a position to judge mechanisms relatively unique to shape completion.

**Fig. 1.**
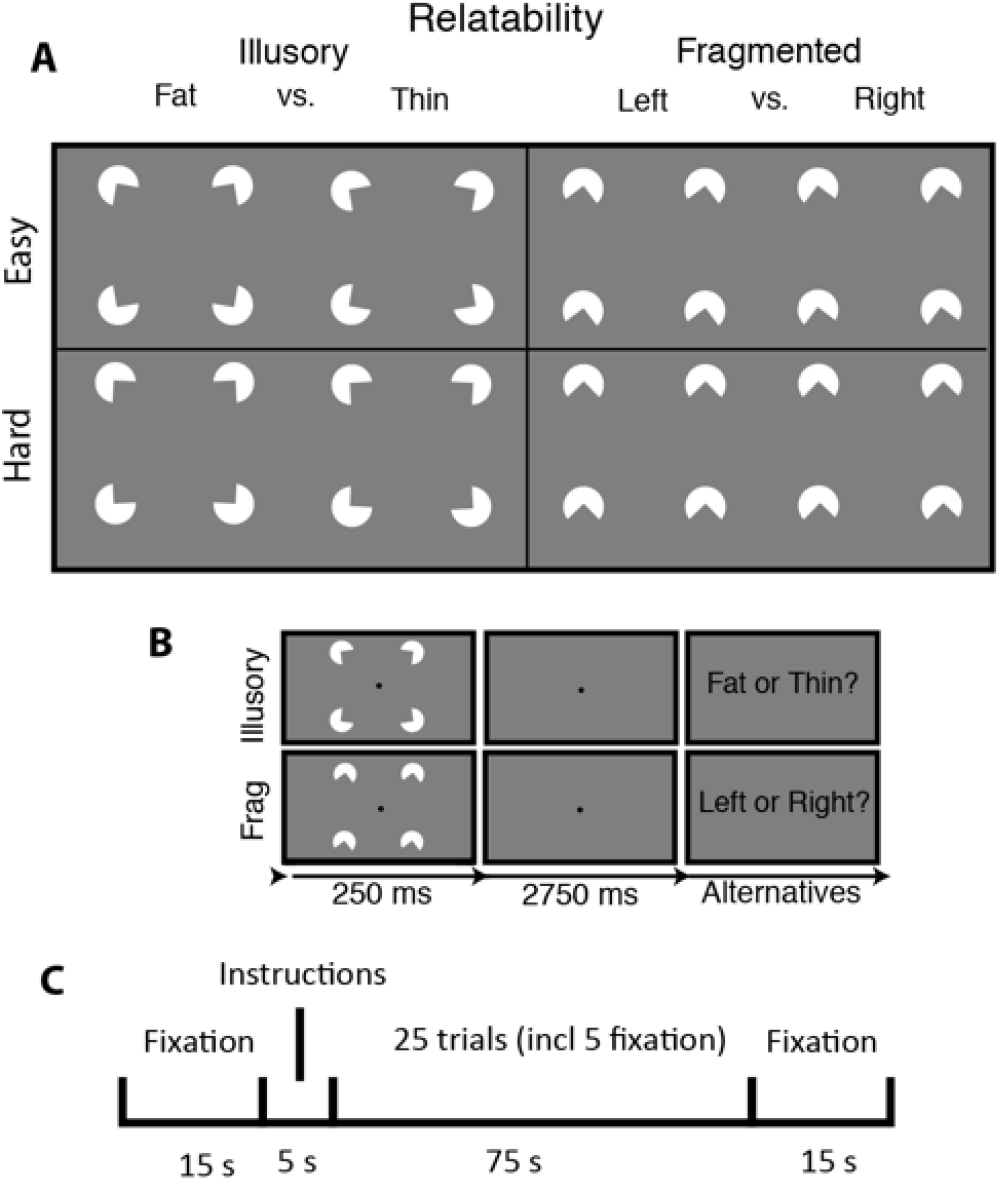
Stimuli, trial sequence, and block arrangement for the visual shape completion experiment. (A) Sectored circles (pac-men) were oriented to generate visually completed shapes (illusory condition) or fragmented configurations that lacked interpolated boundaries (fragmented condition). There were two difficulty conditions corresponding to the amount by which the pac-men were individually rotated to create the response alternatives. (B) After briefly seeing the target, subjects responded with a button press. (C) Each half of a run consisted of a fixation screen, a 5 second instructional screen, 25 trials of a single task condition (including 5 fixation trials), and then another fixation screen. Figure re-used from (Keane et al., 2021a).

Subjects viewed the stimuli in the scanner from a distance of 99 cm by way of a mirror attached to the head coil. There were four white sectored circles (radius = .88 deg, or 60 pixels) centered at the vertices of an invisible square (side = 5.3 deg, or 360 pixels), which itself was centered on a gray screen (see Fig. 1). Stimuli were initially generated with MATLAB and Psychtoolbox code (Pelli, 1997) with anti-aliasing applied for edge artifact removal. Images were subsequently presented in the scanner via PsychoPy (version 1.84; (Peirce, 2007) on a MacBook Pro. Illusory contour formation depended on the geometric property of “relatability” (Kellman and T. Shipley, 1991): when the pac-men were properly aligned (relatable), the illusory contours were present (the “illusory” condition); when misaligned (unrelatable), they were absent (“fragmented” condition).

Within each of the four runs, there was one block of each task condition. Block ordering (illusory/fragmented or vice versa) alternated from one run to the next. Each block had two difficulty levels, corresponding to the magnitude of pac-man rotation (+/- 10 degrees “easy”, or +/- 3 degrees of rotation, “hard”). Within each block, there were 20 task trials and 5 fixation trials. Half of the task trials were easy, and half were hard; half of these two trial types were illusory, and half were fragmented. The ordering of these trial types (including fixation) was counterbalanced. Each trial consisted of a 250 ms pac-man stimulus (task trial) or 250 ms fixation dot (fixation trial), followed by a 2750 ms fixation dot. Subjects needed to issue a response before the end of a task trial; otherwise, a randomly selected response was assigned at the end of that trial and the following trial ensued. Feedback was provided at the end of each run in the form of accuracy averaged cumulatively across all test trials.

Subjects received brief practice outside of and within the scanner before the actual experiment. During practice, subjects were reminded orally and in writing to keep focused on a centrally-appearing fixation point for each trial. To ensure that subjects thoroughly understood the task, pictures of the fat/thin stimuli were shown side-by-side and in alternation so that the differences could be clearly envisaged. Subjects issued responses with a two-button response device that was held on their abdomens with their dominant hand. Subjects practiced with this same type of device outside of the scanner. Feedback after each trial was provided during the practice phase only (correct, incorrect, slow response).

#### 2.3.2. fMRI acquisition

Data were collected at the Rutgers University Brain Imaging Center (RUBIC) on a Siemens Tim Trio scanner. Whole-brain multiband echo-planar imaging (EPI) acquisitions were collected with a 32-channel head coil with TR = 785 ms, TE = 34.8 ms, flip angle = 55°, bandwidth 1894/Hz/Px, in-plane FoV read = 211 mm, 60 slices, 2.4 mm isotropic voxels, with GRAPPA (PAT=2) and multiband acceleration factor 6. Whole-brain high-resolution T1-weighted and T2-weighted anatomical scans were also collected with 0.8 mm isotropic voxels. Spin echo field maps were collected in both the anterior-to-posterior and posterior-to-anterior directions in accordance with the Human Connectome Project preprocessing pipeline (version 3.25.1) (Glasser et al., 2013). After excluding dummy volumes to allow for steady-state magnetization, each experimental functional scan spanned 3 min and 41 s (281 TRs). Scans were collected consecutively with short breaks in between (subjects did not leave the scanner). An additional 10-minute resting-state scan (765 TRs) occurred in a separate session, with the same pulse sequence. Note that due to scanner time constraints one SZ participant finished only 751 of the 765 TRs.

#### 2.3.3. fMRI preprocessing and functional network partition

Preprocessing steps have been reported before (Keane et al., 2021a) but are repeated below. Imaging data were preprocessed using the publicly available Human Connectome Project minimal preprocessing pipeline which included anatomical reconstruction and segmentation; and EPI reconstruction, segmentation, spatial normalization to standard template, intensity normalization, and motion correction (Glasser et al., 2013). All subsequent preprocessing steps and analyses were conducted on CIFTI 64k grayordinate standard space. This was done for the parcellated time series using the Glasser et al. (2016) atlas (i.e., one BOLD time series for each of the 360 cortical parcels, where each parcel averaged over vertices). The Glasser surface-based cortical parcellation combined multiple neuroimaging modalities (i.e., myelin mapping, cortical thickness, task fMRI, and RSFC) to improve confidence in cortical area assignment. The parcellation thus provided a principled way to parse the cortex into manageable number of functionally meaningful units and thereby reduce the number of statistical comparisons. The parcellation also provided units for the brain network partition described further below.

We performed nuisance regression on the minimally preprocessed task data using 24 motion parameters (6 motion parameter estimates, their derivatives, and the squares of each) and the 4 ventricle and 4 white matter parameters (parameter estimates, the derivates, and the squares of each) (Ciric et al., 2017). For the task scans, global signal regression, motion scrubbing, spatial smoothing, and temporal filtering were not used. Each run was individually demeaned and detrended (2 additional regressors per run).

The resting-state scans were preprocessed in the same way as the parcellated task data (including the absence of global signal regression) except that we removed the first five frames and applied motion scrubbing (Power et al., 2012). That is, whenever the framewise displacement for a particular frame exceeded 0.3 mm, we removed that frame, one prior frame, and two subsequent frames (Schultz et al., 2018). Framewise displacement was calculated as the Euclidean distance of the head position in one frame as compared to the one preceding. One HC and one BP did not perform a resting-state scan; one SZ and one BP had too few frames after motion scrubbing (<2.5 standard deviations relative to the mean of their respective subject groups). Group comparisons on the remaining subjects (19 HCs, 15 SZs, and 13 BPs) revealed no differences on either the mean framewise displacement after motion scrubbing (M_HC_=.12, M_BP_=.15, and M_SZ_=.14 mm; F(2,44)=1.49, p=.23) or the mean number of unscrubbed frames (HC—696, BP—632, SZ—663; F(2,44)=1.59, p=.22).

For the task scans, there were 6 task regressors, one for each instructional screen (illusory/fragmented) and one for each of the four trial types (illusory/fragmented, easy/hard). A standard fMRI general linear model (GLM) was fit to task-evoked activity convolved with the SPM canonical hemodynamic response function (using the function spm_hrf.m). Betas for the illusory and fragmented condition were derived from all trials of the relevant condition across all four runs. For the within-group classifier analyses, described below, task activation betas were derived separately for each run, but all other steps were the same as described.

The location and role of each parcel was considered within the context of their functional network affiliations. As noted, an advantage of network-based analyses (rather than individual clusters) is that it substantially increases power to detect average-size effects (Noble et al., 2021). We used the Cole-Anticevic Brain Network partition, which comprised 12 functional networks that were constructed from the above-mentioned parcels and that were defined via a General Louvain community detection algorithm using resting-state data from 337 healthy adults (Ji et al., 2019 see Figure 4A). This partition included: well-known sensory networks—primary visual, secondary visual, auditory, somatosensory; previously identified cognitive networks—frontoparietal, dorsal attention, cingulo-opercular, and default mode; a left-lateralized language network; and three entirely novel networks—posterior multimodal, ventral multimodal, and orbito-affective.

#### 2.3.4. Multivariate pattern analyses

To understand whether specific networks were being used within each subject group, we performed a MVPA with an exhaustive leave-two-runs-out cross-validation for each network (equivalent to split-half cross-validation). This procedure, which has been implemented before (Keane et al., 2021a) and which is applied to each subject individually, entailed determining whether the illusory and fragmented parcel-wise betas for each of the two left-out runs better correlated to the averaged illusory or fragmented betas of the remaining runs (with the number of illusory/fragmented trials always being the same in each run). Similar to past studies, we chose Pearson correlation as the minimum distance classifier (Mill et al., 2020; Mur et al., 2009; Spronk et al., 2020) because it intuitively measures a group’s proximity to an individual in multivariate feature space without requiring parameter choices (e.g., the “C” parameter in support vector machines). Note also that simple linear classifiers perform just as well as sophisticated non-linear methods (e.g., deep learning) with noisy (fMRI) data (Schulz et al., 2020). Results were averaged for each subject across the 6 possible ways to divide the four runs between test and validation. Statistical significance was determined via permutation tests, which generated a null distribution of classification accuracies through the same procedure with 10,000 samples. That is, for each sample and before the cross-validation, the “illusory” and “fragmented” labels were shuffled for each subject and run. The classification results were then averaged across subjects and across the 6 possible divisions of test and validation data sets. False Discovery Rate (FDR) correction was applied to the twelve tests (one for each resting-state networks) (Benjamini and Hochberg, 1995).

To determine which networks were differentially modulated between groups, we conducted, for each pair of groups, a repeated split-half cross-validation using illusory/fragmented activation differences as features. More explicitly, for each repetition of the cross-validation, we considered whether the parcel-wise activation differences (illusory-fragmented) for half of the subjects better correlated with the averaged activation differences of the remaining subjects for each of the two subject groups. Folds were stratified to ensure that each was representative of the overall sample. Results were averaged over 20 repetitions, by which point statistical power plausibly reaches a near-maximum (Valente et al., 2021). Accuracy, sensitivity, specificity, and areas under the curve were calculated using classification values that were averaged across repetitions for each subject. The classifier’s statistical significance was judged relative to a null distribution, which was created by shuffling the subject group labels and repeating the foregoing steps for each of 10,000 samples. Note that the labels were permuted outside of the cross-validation loops, which gives less optimistic (and more realistic) estimates of the underlying null (Etzel and Braver, 2013; Valente et al., 2021). Note also that for each group comparison and across all networks, the mean value of the null distribution always fell near 50% accuracy (range: 49.9-51.2%), demonstrating that sample size imbalances introduced minimal classifier bias. MVPA was applied to each of the twelve resting-state networks and resulting p-values were FDR corrected as before.

#### 2.3.5. Estimating resting-state functional connectivity (RSFC) matrices

For each group, we generated a resting-state functional connectivity (RSFC) matrix to model shape completion via activity flow mapping (see below). We derived each subject’s RSFC by using principal components regression with 100 components, as in past studies (Hearne et al., 2021; Keane et al., 2021a). PC regression was preferred over ordinary least squares to prevent over-fitting (using all components would inevitably capture noise in the data). Multiple regression was preferred over Pearson correlation since the former removes indirect connections (Reid et al., 2019). For example, if there exists a true connection from A to B and B to C, a Pearson correlation, but not regression, would incorrectly show connections between A and C. To generate a subject’s RSFC, for each target parcel time series, we used PCA to decompose the time series of the remaining (N=359) parcels into 100 components, regressed the target onto the PCA scores, and back-transformed the PCA betas into a parcel-wise vector. The average amount of variance explained by the components across subjects was 84% for controls [range: 81-88%], 84% for bipolar patients, [range: 78-89%] and 83% for schizophrenia patients [range: 81-85%]

#### 2.3.6. Brain activity flow mapping (“ActFlow”)

In the next set of analyses, we employed RSFC matrices and brain activity flow mapping to model illusory/fragmented task activation differences. For each subject, task activations in a held-out parcel (‘j’ in Fig. 5A) was predicted as the weighted average of the activations of all other parcels, with the weights being given by the resting-state connections. That is, for each subject, each held-out region’s predicted value was given as the dot product of the task activations in the remaining regions (‘i’ in Fig. 5A) and the subject’s resting-state FC between j and i (using the FC weight from the appropriately oriented regression, i.e., j as the target and i as the predictor). The accuracy of the activity flow predictions was then assessed by computing the overlap (Pearson correlation) between the predicted and actual task activations. Subject-level overlap was expressed by comparing actual and predicted activations for each subject, and then averaging the resulting Fisher-transformed r values (r_z_) across subjects. Statistical significance was determined by comparing the vector of r_z_ values to zero via a one-sample t-test. ActFlow has yielded accurate estimates of task-evoked activations for cognitive control, visual working memory, and visual shape completion tasks, among others (Cole et al., 2016; Hearne et al., 2021; Keane et al., 2021a).

We applied the ActFlow methodology to consider possible group differences in how other networks interfaced with the secondary visual network. The secondary visual network was of interest because it is central to shape completion in healthy adults (Keane et al., 2021a) and because particular regions falling within this network (i.e., LO, V4) have been repeatedly implicated in shape completion via EEG, MEG, TMS, and single-unit recording (Cox et al., 2013; Halgren et al., 2003; Murray et al., 2006; Wokke et al., 2013). We considered how ActFlow estimates improved in that network, when any of the remaining networks were individually added (Fig. 3). This change was determined simply by comparing via a paired t-test the prediction accuracies (Fisher Z-transformed correlations) before and after adding each network. A significant improvement would indicate which other networks, if any, guided activity flow in the secondary visual network.

**Fig. 2.**
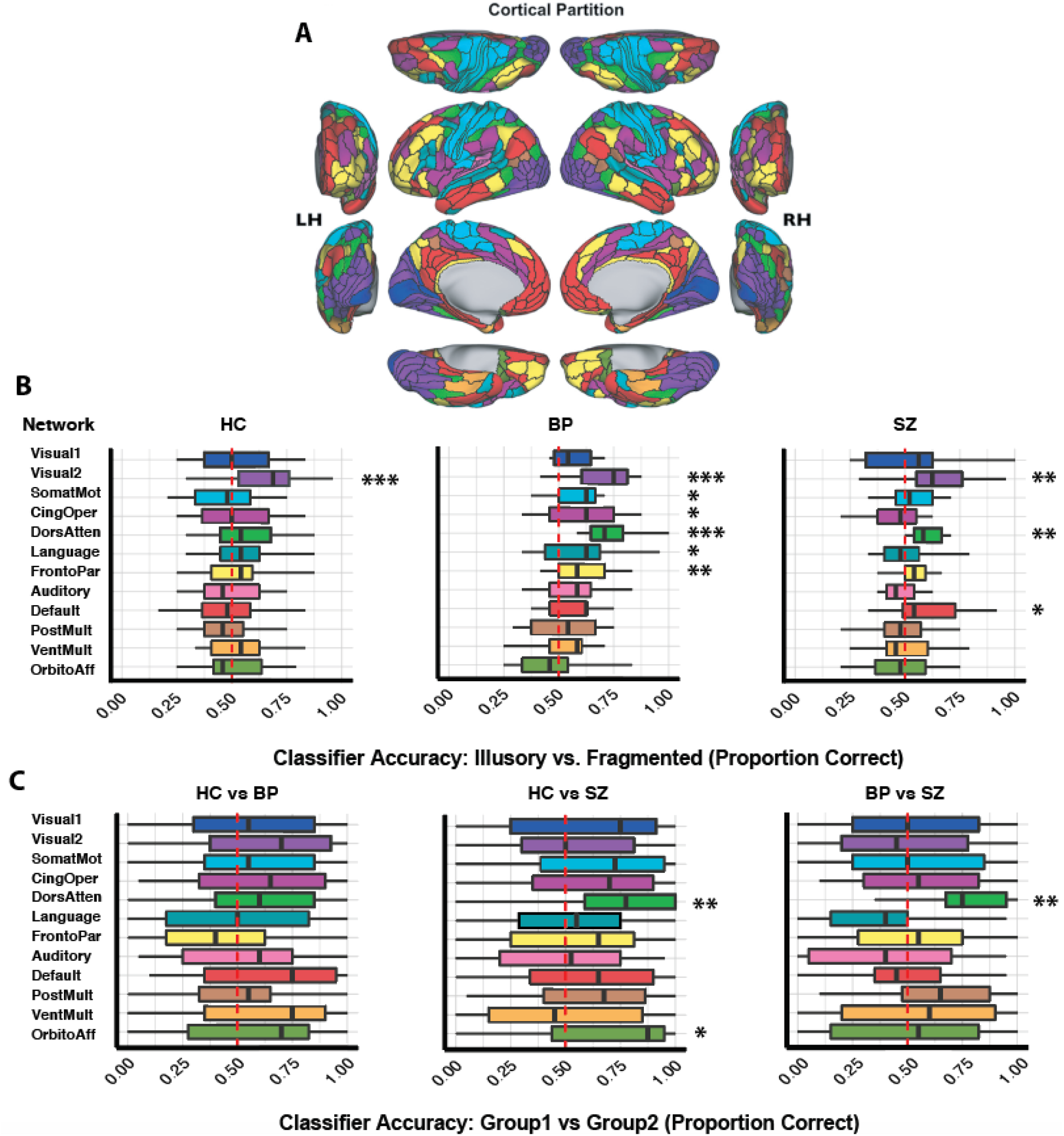
(A) The twelve functional networks (Ji et al., 2019) are color-coded to match panels B and C. (B) Box plots depicting illusory/fragmented classification accuracy for each group using leave-two-runs-out cross-validation. The red dotted lines demarcate 50% performance. (C) Box plots depicting group classification accuracy for each pair of groups using repeated split-half cross-validation, where the features correspond to illusory/fragmented differences. See text for additional classification statistics. *p_corr_<.05, **p_corr_<.01, ***p_corr_ <=.001.

**Fig. 3.**
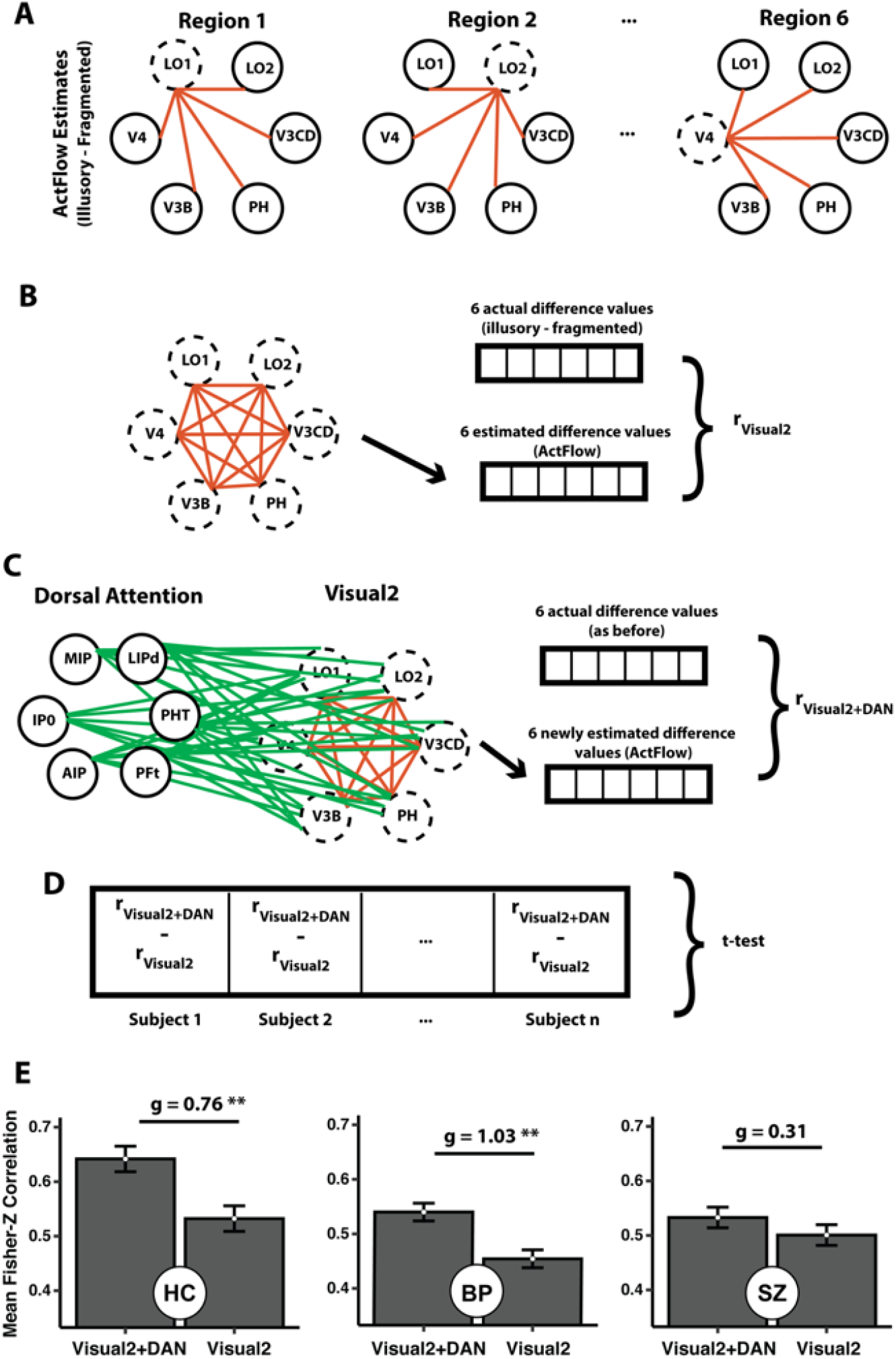
Gauging modeling contributions of the dorsal attention network (DAN) to the secondary visual network (Visual2). (A) For a given subject, task activation differences for each significant Visual2 parcel were estimated (dotted circles) using *actual* task activation differences in the remaining parcels (solid circles) and their resting-state connections (red lines). For illustration, only six regions are shown for each network. (B) ActFlow accuracy was defined as the correlation between actual and estimated task activation differences, across the Visual2 parcels. (C) Task activation differences were again estimated via ActFlow, except that, this time, the connections and activation differences from dorsal attention regions could also contribute. (D) The difference between the original and re-calculated estimates was computed for each subject (after a Fisher Z-transform) and directly compared across subjects within a group via a one-sample (or paired) t-test. (E) The DAN could significantly improve ActFlow estimates in the secondary visual network in controls and bipolar patients, but not in schizophrenia patients. Errors=+/-SEM **p<.01. Figure is adapted from Keane et al. (2021a).

#### 2.3.7. Predicting cognitive disorganization from illusory/fragmented parcel-wise modulations

The DAN was differentially modulated in SZs relative to the other groups in the aforementioned MVPA. Cognitive disorganization has been associated with impaired shape completion and altered neural oscillations as noted (Keane et al., 2019; Spencer et al., 2004; Spencer and Ghorashi, 2014). Can these results be linked more directly? To consider the question, we ran a linear regression with leave-one-out cross-validation. Leave-one-out was chosen because, contrary to popular conceptions, it generally yields the least bias/variance for prediction (Zhang and Yang, 2015) and because its predictions can generalize surprisingly well to held-out fMRI data (Anticevic et al., 2014; Rosenberg et al., 2015). Within each training loop, the outcome variable (disorganization) and each predictor variable (modulations for a given DAN parcel) were z-scored using the means and standard deviations from the training set (to prevent circularity) (Mill et al., 2020; Shen et al., 2017). In the training set, the disorganization scores were regressed onto the modulations and the resulting beta coefficients were used to predict the held-out subject’s disorganization score. Model prediction accuracy was gauged as the mean absolute error between predicted and actual disorganization (MAE). Statistical significance was judged via permutation testing. That is, we compared the true MAE to a distribution of such values that were generated by randomly shuffling the disorganization scores across subjects (without changing the feature matrix). As before, the disorganization variable was reshuffled once for each of the 10,000 samples of the null distribution, before the cross-validation loops. To demonstrate robustness, we additionally ran repeated leave-two-out and 10-fold cross-validation. The method was the same as just described except that MAE was averaged across 100 randomized splits between test and training.

#### 2.3.8. Experimental design and statistical analysis

Analyses were performed with RStudio (Version 1.2.1335) and MATLAB R2019a, except for the behavioral analyses which were done with SPSS version 27. Cortical visualizations were created with Workbench (version 1.2.3). Between-group variance was removed from error bars when reporting the standard error of the means in within-subject comparisons (Loftus, 1994). The final sample sizes were determined by the duration of funding (see Acknowledgements and section 2.1 above). False Discovery Rate correction, when applied, was denoted by p_corr_ and had a threshold of q<.05 (Benjamini and Hochberg, 1995). T-test effect sizes were given as Hedges’ g and were generated with the measures-of-effect-size toolbox in MATLAB (Hentschke, 2021).

#### 2.3.9. Data/code accessibility

Brain activity flow mapping MATLAB code is part of the freely-available ActFlow toolbox: https://github.com/ColeLab/ActflowToolbox. HCP minimal preprocessing pipelines are also publicly available: https://github.com/Washington-University/HCPpipelines/releases. The Cole Anticevic Brain Network partition can be found here: https://github.com/ColeLab/ColeAnticevicNetPartition. Neural data will be released on OpenNeuro.org along with resting-state functional connectivity matrices and unthresholded task activation maps.

## 3. Results

### 3.1. Behavioral task performance

Employing a 2 (task condition) by 2 (difficulty) by 3 (group) within-subjects ANOVA (type III sum of squares), we found that performance was more accurate in the fragmented than illusory condition (88.9% versus 80.9%, F(1,48)=28.9, p<.01) and better in the (“easy”) large-rotation condition than the “hard” small-rotation condition (F(1,48)=229.5, p<10^−19^) (See Table 2). The accuracy difference between illusory and fragmented conditions did not depend on difficulty level, although there was a trend toward a greater difference on the smaller rotation condition (two-way interaction: F(2,48)=3.3, p=.08). The marginal interaction probably arose from ceiling effects for the fragmented condition since there was no corresponding interaction in the reaction time data (F(2,48)=.82, p=.37). Reaction time data were in other ways entirely predictable from the accuracy results, with faster performance in the fragmented than the illusory condition (F(1,48)=11.4, p<.01), and faster performance in the easy than the hard condition (F(1,48)=65.7, p<10^−9^). The no-response trials were infrequent, occurring on only 3.9% of the trials on average. The frequency of no-response trials did not vary with difficulty level or task condition nor was there an interaction between difficulty and task condition (all p>.10). Note that one SZ patient exhibited chance task performance but was retained so as to have a more typical and representative patient sample. Most importantly, across all three ANOVAs (accuracy, RT, no-response frequency), there were no main effects or interactions with subject group (all p>.28; all partial eta squared<.055). Note that we were not necessarily expecting significant behavioral differences with our sample sizes since the group difference in a larger-scale study was of medium-large magnitude (d=.67; 134 patients, 66 HCs) (Keane et al., 2019). We nevertheless propose reasons in the Discussion why our observed shape completion deficits were smaller than anticipated.

**Table 2.** Task Performance.

|  | HC | BP | SZ |
| --- | --- | --- | --- |
| % Correct, Illus | 82.9 (3.0) | 80.0 (3.5) | 79.6 (3.4) |
| % Correct, Frag | 89.6 (2.2) | 89.7 (2.5) | 87.4 (2.4) |
| Reaction Time (s), Illus | 1.04 (.07) | .93 (.08) | 1.06 (.07) |
| Reaction Time (s), Frag | .94 (.07) | .84 (.08) | .99 (.08) |
| % Slow Response, Illus | 4.3 (2.6) | 0.4 (3.0) | 5.2 (2.9) |
| % Slow Response, Frag | 6.8 (2.8) | 0.6 (3.2) | 6.3 (3.1) |

*Note. Mean values for each variable (with standard error of the mean)*

Consistent with past results (Keane et al., 2021a; 2019), the fragmented and illusory conditions were highly correlated behaviorally across subjects (accuracy—r=.67, p<10^−7^; RT— r=.85, p<.10^−11^), confirming that the two were reliant upon a common core of mechanisms. The correlations were robust and remained significant when calculated with non-parametric tests or after log-transforming the RT data.

### 3.2. Traces of shape completion were scattered over more networks in patients

To determine the networks relevant to shape completion in each group, we ran a leave-two-runs-out MVPA, which assessed—for each subject and network—whether the illusory and fragmented betas from the training runs could be used to correctly classify the illusory and fragmented betas from the two remaining runs. To determine the involvement of each network within a group, classification accuracy results were aggregated across subjects and compared to a null distribution (see Methods). For healthy controls, the secondary visual network encoded the modulations, as already reported (accuracy=63%, p_corr_=.001) (Keane et al., 2021a). For schizophrenia patients, three networks encoded the modulations: secondary visual (accuracy=63%, p_corr_=.002), dorsal attention (accuracy=61%, p_corr_=.008), and default mode (accuracy=59%, p_corr_<.05). Finally, for bipolar patients, *six* significant networks encoded the modulations: secondary visual (accuracy=69%, p_corr_<.0001; somatomotor (accuracy=59%, p_corr_=.03), cingulo-opercular (accuracy=60%, p_corr_=.01), dorsal attention (accuracy=71%, p_corr_<.0001), language (accuracy=59%, p_corr_=.03), and frontoparietal (accuracy=59%, p_corr_=.010). These results suggest that whereas the secondary visual network was important in each subject group (essentially an out-of-sample replication of the control group result) additional cognitive networks also appeared relevant. These results also show that multivariate traces of shape completion were distributed through more networks in patients.

### 3.3. Dorsal attention and orbito-affective networks distinguished schizophrenia patients

To consider whether groups could be distinguished in parcel-wise patterns, we trained MVPA classifiers separately for the 12 functional networks (Ji et al., 2019). For each pair of subject groups and for each network, the classifier used illusory/fragmented activation differences to categorize subjects by their group membership (see Methods). After FDR correction, no network could distinguish bipolar patients and healthy controls. The networks that could distinguish schizophrenia patients from healthy controls were the dorsal attention (p_corr_ = .005, sensitivity_SZ_ = .74, specificity_SZ_ = .74, AUC = .86) and orbito-affective (p_corr_ = .03, sensitivity_SZ_=.65, specificity_SZ_ = .72, AUC = .77). When comparing bipolar and schizophrenia patients, only the DAN reached significance (p_corr_=.007, sensitivity_SZ_=.73, specificity_SZ_=.73, AUC=.87). The secondary visual network did not distinguish any of the three groups (all p_corr_>.24). To summarize, patterns of dorsal attention task activations could distinguish schizophrenia patients from the other groups; and the orbito-affective network was also able to distinguish SZs from controls. These results, which will be elaborated upon more fully in the Discussion, are consistent with our hypothesis that brain network differences during visual perceptual organization in schizophrenia are primarily related to higher-level cognition.

### 3.4. Dorsal attention network activity was related to cognitive disorganization

Increased cognitive disorganization has been associated with poorer shape completion and abnormal oscillations, as noted (Keane et al., 2019; Spencer et al., 2004; Spencer and Ghorashi, 2014). Task-related DAN modulations can distinguish SZ patients from other subject groups (Fig. 3C). Can these variables be linked more directly? To consider the question, we individually regressed each clinical variable onto the modulations of the 23 dorsal attention parcels using leave-one-out cross-validation with permutation testing (see Methods). Across all 31 patients, the modulations were indeed related to cognitive disorganization (r=.65, MAE=.78, p=.007). These results were robust and would also be significant if we were to use leave-two-out or 10-fold cross-validation (both p<=.01).

### 3.5. Potentially reduced feedback activity from dorsal attention to secondary visual networks in schizophrenia

A recently-developed predictive modeling approach—activity flow mapping (“ActFlow”) (Cole et al., 2016)—has demonstrated that resting-state connections are likely relevant to shape completion in healthy controls (Keane et al., 2021a). This method computes the activation difference (illusory minus fragmented) in a held-out “target” parcel as the linear weighted sum of the activation differences in all other parcels, with the weights being given by the resting-state connections to the target (Fig. S2 in Supplementary Materials). This algorithm is based on neural network simulations, and can thus be thought of as a rough simulation of the movement of task-evoked activity that likely contributed to each brain region’s task-evoked activity level, which in turn can provide evidence that the resting-state connections mechanistically support shape completion. As described in more detail in the Supplementary material, when applied to all 360 parcels across cortex, the ActFlow modeling generated accurate results for all three groups (all r>.56, all p<10^−6^) and this accuracy did not differ between groups (all p>.23).

We developed a novel extension of the ActFlow framework, which shows that—in healthy controls—the DAN can model activity in the secondary network, potentially reflecting feedback to mid-level visual areas (Keane et al., 2021a). In this method, we computed a single correlation between the actual and estimated parcel difference values (illusory-fragmented) across the 54 secondary visual network parcels. We then recomputed this correlation, when each of the 54 parcels could also be predicted by parcels and connections from the 23 dorsal attention regions (see Fig. 3). Finally, we Fisher-z transformed the correlations, subtracted the two, and then performed a one-sample t-test to see if the correlations increased as a result of the network’s inclusion. Can the DAN model activity in the secondary visual network in the patient groups? As shown in Fig. 3E, the DAN improved the predictions for the secondary visual network in healthy controls (Δr≈Δr_Z_=.11; *t*(18)=3.3, *p*=.004, *g*=.76) and bipolar patients (Δr≈Δr_Z_=.09; *t*(12)=3.7, *p*=.003, *g*=1.03), but not in schizophrenia patients (Δr≈Δr_Z_=.03; *t*(14)=1.2, *p*=.25, *g*=.31). No other network could model the secondary visual network in any group. These results would be the same with an FDR correction (across all networks). Thus, DAN fails to robustly model the secondary visual network in schizophrenia, perhaps because of reduced feedback from dorsal attention to secondary visual areas.

## 4. Discussion

Visual shape completion plays a critical role in extracting object shape, size, position, and number from edge elements dispersed across the field of view. Prior electrophysiological and psychophysical work has suggested that schizophrenia patients properly form illusory contours at initial stages of processing but potentially exhibit later-stage differences related to cognitive control. However, these findings have not been corroborated with other neuroscience methods and were generally limited by their lower spatial resolution. Here, we leveraged recent tools in computational neuroimaging to investigate the functional connections and brain networks that may differ in schizophrenia during shape completion. We additionally considered whether such differences arise in bipolar disorder or whether they vary monotonically with illness factors that cut across the schizo-bipolar spectrum. It was hypothesized that cognitive—but not visual—networks would be differentially active in SZ, that activity in one of these networks would be linked to cognitive disorganization, and, more speculatively, that top-down feedback to the secondary visual network would be faulty in SZ.

Five major findings emerged. First, the DAN was differentially active in SZ compared to the other groups. Next, the secondary visual network was strongly modulated within each group and did not differ group-wise in its activation pattern. Third, dorsal attention modulations across all patients were related to cognitive disorganization severity. Fourth, in schizophrenia, our modelling showed little influence of the DAN on the secondary visual network (in contrast to the other groups, who did show an influence). A final unanticipated finding was that— regardless of diagnosis—patients incorporated more networks overall during shape completion. Below, we discuss these findings in more detail, identify potential limitations, and suggest future directions along the way.

### 4.1. Aberrant DAN activity in schizophrenia

Dorsal attention network activity was unequivocally aberrant in SZ. This is consistent with earlier SZ studies, which have argued for abnormal dorsal stream contributions to motion perception (O’Donnell et al., 1996), stereopsis (Schechter et al., 2006), and fragmented object recognition (Sehatpour et al., 2010). Visual working memory deficits (and broader indices of cognition) have also been attributed to abnormal activation in posterior parietal cortex (Hahn et al., 2018), which overlaps with the DAN. A goal for future research will be to determine to what extent abnormal DAN activity emerges across these and other visual tasks in SZ. Because DAN differences were so large and because they were found relative to both healthy controls and bipolar disorder patients (AUCs>.85; sensitivities>.72; specificities>.72), such differences might yield a candidate biomarker for differential diagnosis or predicting future psychosis onset. More highly powered studies with early-stage or high-risk patients are needed to confirm these possibilities.

DAN task activity was also related to a central feature of psychosis, cognitive disorganization. The brain networks undergirding cognitive disorganization—alternatively referred to as “formal thought disorder” or “conceptual disorganization”—are largely unknown perhaps because disorganization is less often parsed out as a separate construct (with many studies preferring instead to lump it in with the more encompassing positive symptom factor). An interesting possibility going forward will be to examine whether DAN activity during perceptual organization can serve as a proxy for cognitive disorganization or whether stimulating parts of DAN can ameliorate symptom severity.

### 4.2. Possibly reduced top-down attentional feedback in schizophrenia and more “cognitive” perceptual organization in SZ

It is not possible to tease apart feedforward and feedback activity using the hemodynamic response. However, a realistic possibility is that DAN dysfunction may disrupt top-down attentional amplification (Berkovitch et al., 2017; 2018), which may be needed to properly notice and use illusory contours for shape discrimination (Keane et al., 2012). Such disruption has been linked specifically to cognitive disorganization, NMDA receptor hypofunction, and gamma band synchrony abnormalities, all of which characterize the schizophrenia phenotype (Berkovitch et al., 2017). This view fits with past behavioral work showing that SZ patients have impaired top-down attentional control for noticing subtle stimuli (Gold et al., 2007; Luck and Gold, 2008); it also fits with the assertion that people with schizophrenia have impaired dorsal top-down feedback to ventral object-recognition areas (Tapia and Breitmeyer, 2011).

In light of the significant DAN activity in the SZ group, we speculate that patients may compensate for poor top-down modulation by carrying out computations within the DAN itself or by the DAN interfacing with other non-visual networks. This view of perceptual organization as being more cognitively reliant might also explain why conceptual knowledge aids interpretation of a vague visual stimulus more for people with psychosis or psychotic-like experiences than for people without such conditions (Teufel et al., 2015).

The above sketch implies that perceptual organization deficits may become more prominent if the (longer-latency) cognitive networks are given less time to operate (Wyatte et al., 2014). The current study presented a 250 ms pac-man configuration with no mask (to ensure a more robust BOLD response), but earlier psychophysical studies presented the pac-men for 200 ms with a mask 50 ms afterwards (Keane et al., 2021b; 2019). This difference may help explain why the shape completion in a prior larger-scale study was medium-to-large (d=.67) (Keane et al., 2019), whereas the group difference in the current study was small (d=.10). A prediction is that if we were to present stimuli more briefly with shorter masking SOAs or with more disruptive masks, then shape completion deficits should emerge more clearly. Another prediction is that applying single-pulse transcranial magnetic stimulation over dorsal attention regions 200-300 ms after stimulus onset (Wyatte et al., 2014) may elicit stronger shape completion impairments in schizophrenia than in controls.

### 4.3. Orbito-affective dysfunction in SZ during perceptual organization

Modulations in the recently-defined orbito-affective network could distinguish SZs and HCs and this too suggests that higher-order cognition may play a more important role for patients. We had speculated in a past psychophysical investigation that orbitofrontal cortex could be relevant (Keane et al., 2014) because: i) this region is differentially active 300 ms after stimulus onset during Kanizsa shape detection tasks in healthy adults (Halgren et al., 2003); ii) the orbitofrontal cortex contributes to object recognition under impoverished viewing conditions (Bar, 2003); and iii) gray matter volume in the region gradually shrinks over the course of the illness, especially among persons with thought disorder (Nakamura et al., 2008). The orbito-affective network overlaps with posterior orbitofrontal cortex, which is associated with “reward processing” (Kahnt et al., 2010) and is strongly connected to the ventral striatum, the substantia nigra/ventral tegmental area, and the globus pallidus (Ji et al., 2019). Some of these same regions are routinely found to be hyperconnected to visual cortex in SZ (Anticevic et al., 2014; Damaraju et al., 2014) and are associated with dopaminergic dysregulation in in unmedicated schizophrenia patients (Horga et al., 2016). While it is outside the scope of this work, it is worth investigating the cooperation between these subcortical structures and the orbitofrontal cortex as SZ patients attempt to recognize partly visible shapes.

### 4.4. Bipolar disorder and schizophrenia have more diffuse neural representations

An unexpected but interesting finding was that completed shapes were encoded across a broader range of networks in each disorder. The reason for this more distributed neural representation is unknown but could be because computations ordinarily performed by vision could have been outsourced to nominally non-visual networks as suggested above; or they could be byproducts of a less modularized brain network architecture (Ma et al., 2020). Networks in bipolar disorder may be especially less well-integrated and less centralized as compared to those in healthy controls or even schizophrenia (van Dellen et al., 2020). It is worth considering in future research whether similarly diffuse neural representational patterns emerge in other visual and cognitive tasks.

### 4.5. Addressing limitations

The most obvious limitation is sample size. However, we avoided stringent multiple comparison corrections by restricting our analyses to only 12 pre-defined networks. Network-based analyses are also plausibly more powerful in that they pool weaker parcel-wise effects over larger functionally related areas (Cremers et al., 2017; Ji et al., 2019; Noble et al., 2021). Moreover, the implicated networks in our analysis did not emerge out of the blue but were suspected on the basis of past psychophysical and electrophysiological work (see Introduction). Additionally, the abnormal DAN activity in SZs was shown relative to two clinically and demographically well-matched groups (Table 1); both effects were large and could survive an FDR (or a Bonferroni) correction. Finally, the cognitive disorganization relation was anticipated from past work (Keane et al., 2019; Spencer et al., 2004; Spencer and Ghorashi, 2014) and was shown with multiple forms of cross-validation using the larger combined patient sample (n=31).

Another limitation is that patients were on medication. Note, however, that the patient groups did not significantly differ on olanzapine equivalents and prior behavioral and electrophysiological studies found no relationships between shape completion and the type/dose of neuroleptics (Foxe et al., 2005; Keane et al., 2019; Spencer and Ghorashi, 2014).

Another objection is that groups differed in their eye movements and this may have confounded the results. This too is unlikely since: 1) pac-men locations were equidistant from fixation, equally informative within a trial, and matched between conditions, reducing the chance of systematic task condition differences; 2) the illusory and fragmented conditions were highly correlated in RT and accuracy and groups were undifferentiated on RT and accuracy, suggesting again that any possible eye movement differences impacted performance minimally; 3) saccading after stimulus onset would offer little benefit since saccade latency is ∼200 ms (Sumner, 2011) and the stimuli appeared for only 250 ms at unpredictable times during a block (See also Keane et al. (2021a); 4) there is little evidence that eye movements impact visual shape completion in non-translating displays and some evidence that it has no effect relative to a control “fragmented” condition (Cox et al., 2013 see the fixational heat maps in their Figure S2).

A related objection is that subjects covertly attended to—and responded on the basis of—exactly one pac-man. Aside from being contrary to task demands, this again is unlikely since it is doubtful that any one network would be differentially modulated in such a scenario. The fact that the secondary visual network could strongly differentiate the two conditions, consistent with past research (Keane et al., 2021a), suggests that each group represented completed shapes in one condition but not in the other.

There could also be residual confounds: the Glasser atlas could have been inappropriate for patient groups (e.g., due to differences in cortical folding), lithium intake could have made the neural results incommensurable, or imperfect motion correction could have still led to group differences. However, RSFC matrices were similar between groups on univariate, multivariate, and Mantel tests, arguing against such confounds (see Supplementary materials).

To summarize, employing a well-validated perceptual organization task, we revealed clinically relevant DAN abnormalities in schizophrenia, orbitofrontal differences in schizophrenia, and more distributed shape representations across all patients, potentially reflecting compensatory mechanisms. Goals for future research will be to establish a causal role for the DAN and to consider whether briefer (masked) stimulus presentations can minimize potential compensatory cognitive influence or generate more obvious group differences in shape completion performance.

## Data Availability

Neural data will be released on OpenNeuro.org along with resting-state functional connectivity matrices and unthresholded task activation maps.

## Acknowledgments

We thank Rebekah Boy, Laura Crespo, Lisa Cruz, Blair Singer, and Dillon Smith for help in recruiting participants and collecting and organizing study data. We are also indebted to Michael Harms for assistance in finalizing the pulse sequence, Pamela Butler for assistance in patient recruitment, and Takuya Ito and Carrisa Cocuzza for providing sample code. The authors additionally acknowledge the Office of Advanced Research Computing (OARC) at Rutgers University for providing access to the Amarel cluster and associated research computing resources (http://oarc.rutgers.edu). This work was supported by a National Institutes of Health Mentored Career Development Award (K01MH108783) to BPK.

## Supplementary Methods and Results

### Similar RSFC matrices between groups

Group differences/similarities in RSFC were assessed in three ways. First, we compared groups on each connection weight in the full 360×360 matrix using FDR-corrected independent t-tests. After FDR correction, there were no significant differences on any pairwise comparisons. Second, we considered whether *patterns* of connection weights differed between groups, by vectorizing the lower triangle of each subject’s RSFC matrix and employing the between-group MVPA method described above in section 2.3.4. This was done for each of the 12 resting-state networks, with FDR-correction as before. There were no significant differences between any pair of groups for any network (all p_corr_>.27). Finally, we quantified the *similarity* between FC matrices via the non-parametric Mantel permutation test (Mantel, 1967; Spronk et al., 2020), which accounts for the non-independence of FC matrix values (Diniz-Filho et al., 2013). This test was conducted by i) taking the lower triangle of the RSFC matrices of each subject, ii) averaging the vectorized regression weights element wise across subjects within group, and iii) computing a Pearson’s R between the two group-averaged vectors. Statistical significance of the resulting correlation was judged relative to a null distribution, which was generated the same way except that the region identities were shuffled for each of 10,000 samples. As in prior work, the resulting null distribution for this RSFC analysis was converted into a probability distribution function (using MATLAB function ksdensity) before calculating a p-value (Spronk et al., 2020). We found similar RSFC for each pair of groups for the 360 × 360 matrix (HC vs. BP: r=.76, p<10^−7^ ; HC vs. SZ: r=.77, p<10^−7^; BP vs. HC: r.=.73, p<10^−7^). As an additional comparison, and for illustration purposes only, we also compared the RSFC of the regions that were significantly modulated during the task (illusory-fragmented) among healthy controls (Keane et al., 2021a). Conducting the same analyses as just described, there were no significant differences, after correcting for multiple comparisons. The broadly similar RSFC matrices are consistent with past results (Spronk et al., 2020) and provide evidence that each group’s data were of similar quality.

**Fig. S1.**
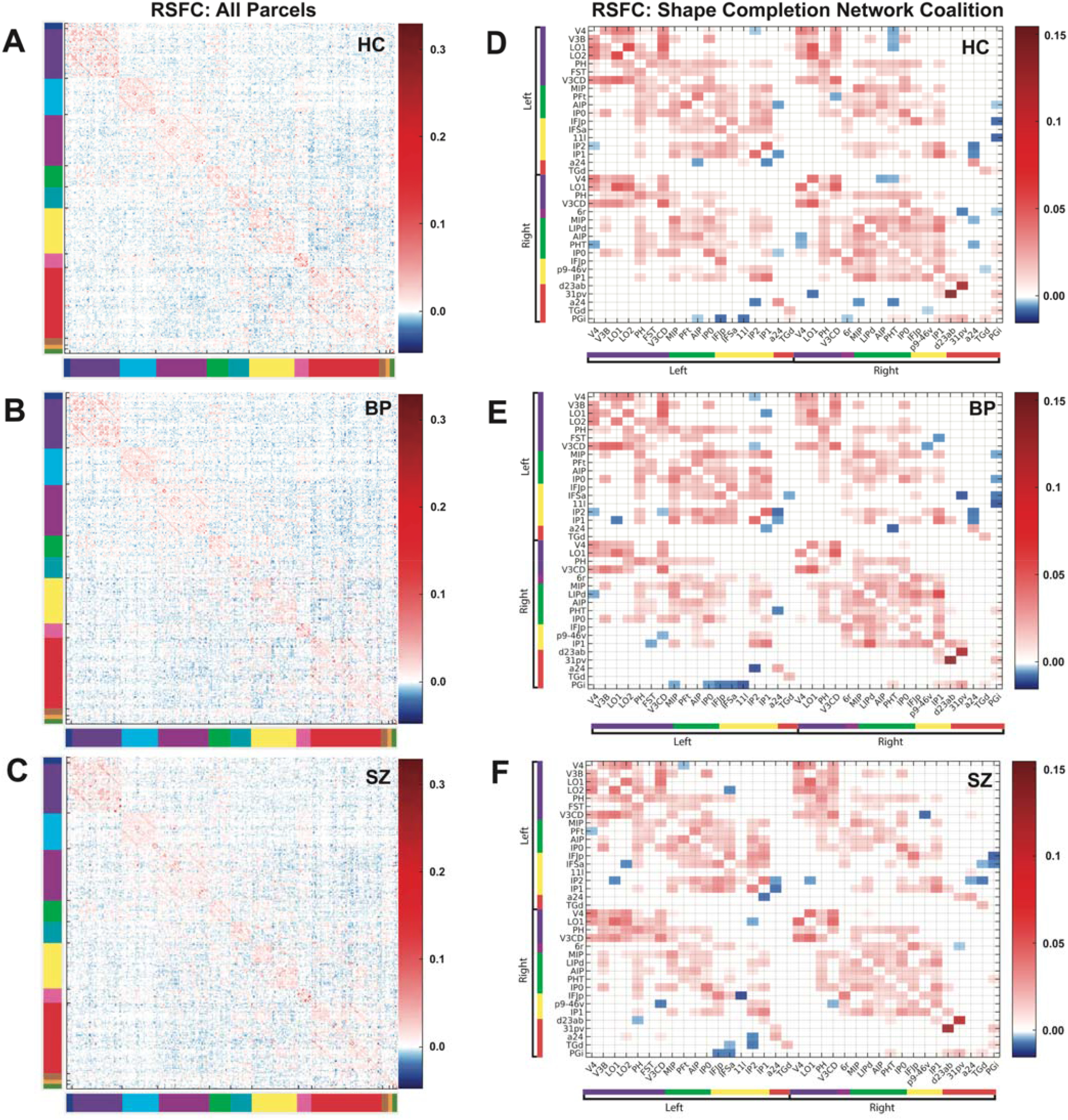
Resting-state functional connectivity (RSFC) matrices were similar for each subject group. (A-C) A full unthresholded 360×360 RSFC matrix showing group-averaged regression coefficients for healthy controls, bipolar disorder patients, and schizophrenia patients. The blue/red colors indicate the degree to which a given parcel time series was predicted by all remaining parcels. Colors on the outskirts of the matrices indicate the network to which a given region belonged (see Fig. 2A). (D-F) As an additional comparison, we also compared the functional connectivity matrices using the three dozen regions that were significantly task modulated among healthy controls (using a univariate illusory-fragmented contrast). As can be seen, the RSFC matrices were very similar. Each trio of matrices was scaled in the same way across groups to enable cross-group comparisons.

### Resting-state connections likely subserve shape completion in each subject group

A recently-developed predictive modeling approach—activity flow mapping (“ActFlow”) (Cole et al., 2016)—has demonstrated that resting-state connections are likely relevant to shape completion in healthy controls (Keane et al., 2021a). This method computes the activation difference (illusory minus fragmented) in a held-out “target” parcel as the linear weighted sum of the activation differences in all other parcels, with the weights being given by the resting-state connections to the target (Fig. S2). This algorithm is based on neural network simulations, and can thus be thought of as a rough simulation of the movement of task-evoked activity that likely contributed to each brain region’s task-evoked activity level, which in turn can provide evidence that the resting-state connections mechanistically supported shape completion. Prediction accuracies (correlations between the actual and predicted activation differences computed for each individual subject) were well above zero at the whole-cortex level for each group (HC: *r*=.64, *p*<10^−9^; BP: r=.63, p<10^−8^; SZ: r=.57, p<10^−6^). Between-group comparisons of these correlations (with Fisher-Z transforms) yielded no significant differences (F(2,44)=.9, p=.42; eta squared=.04; on pairwise comparisons, all g <.41 and all p>.23 uncorrected). Taken together, these results demonstrate that the resting-state connectivity data could be used to model shape completion activations in each group. These results attest to the appropriateness of the brain activity flow mapping procedure for understanding shape completion in healthy and schizo-bipolar populations and lend credibility to the modeling results in the main article.

**Fig. S2.**
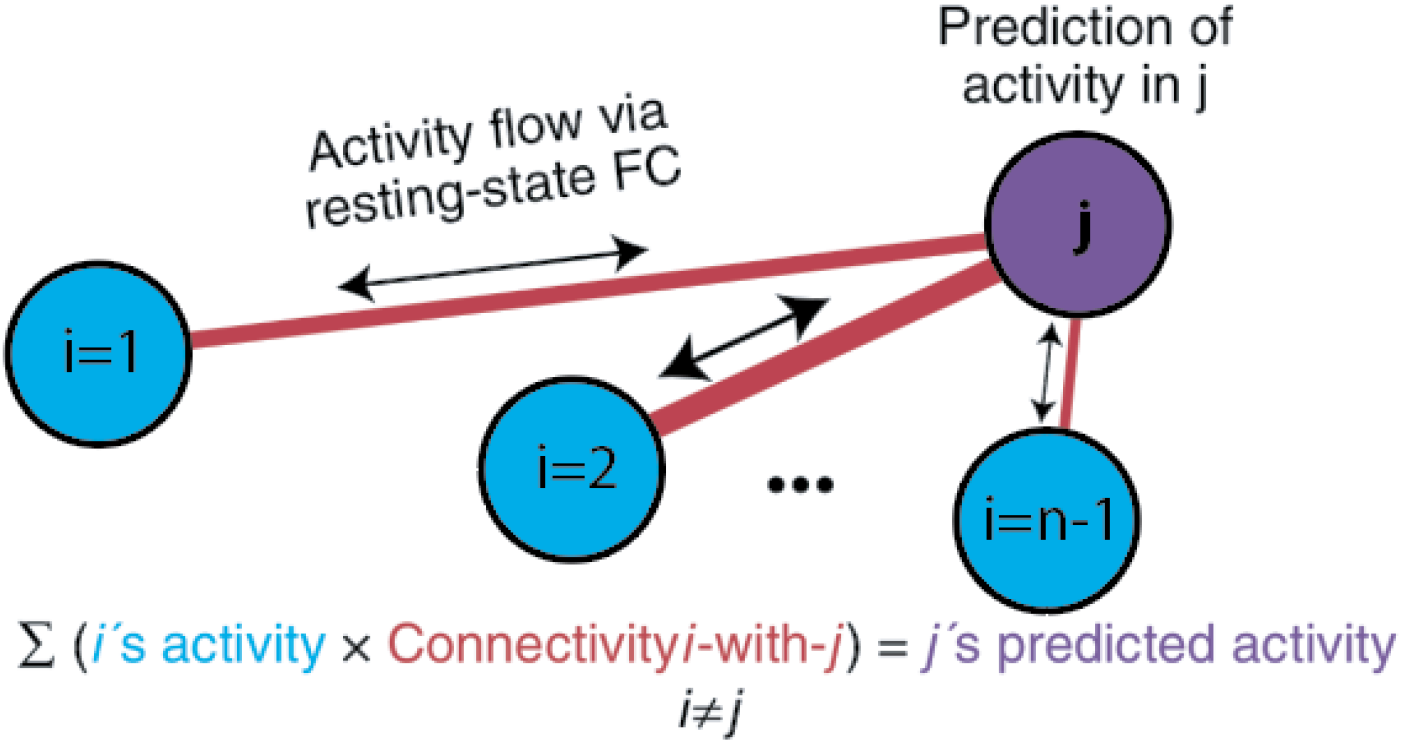
Activity flow mapping procedure. For each subject, the task activation differences (illusory-fragmented) in a held-out parcel (j) is given by the dot product between the activation differences in the remaining parcels (regions i) and the resting-state connection strengths (betas) between i and j. Thicker red lines denote stronger functional connections.

## Notes

*Declarations of Interest*: The authors declare no competing conflicts of interest.

### Competing Interest Statement

The authors have declared no competing interest.

### Funding Statement

This work was funded by a National Institutes of Health Mentored Career Development Award (K01MH108783) to BPK.

### Author Declarations

The study followed the tenets of the Declaration of Helsinki and was approved by the Rutgers University Institutional Review Board.

### Summary of Updates

The manuscript has been shortened and the introduction and discussion have been rewritten. Certain analyses have even placed in the 'Supplementary material' near the bottom of the manuscript.

